# Mutational signatures driven by epigenetic determinants stratify patients for therapeutic interventions in gastric cancer

**DOI:** 10.1101/2020.04.17.20066944

**Authors:** Jaqueline Ramalho Buttura, Monize Nakamoto Provisor, Renan Valieris, Vinicius Fernando Calsavara, Rodrigo Duarte Drummond, Alexandre Defelicibus, Joao Paulo Lima, Helano Carioca Freitas, Vladmir C. Cordeiro Lima, Thais Fernanda Bartelli, Marc Wiedner, Rafael Rosales, Kenneth John Gollob, Joanna Loizou, Emmanuel Dias-Neto, Diana Noronha Nunes, Israel Tojal da Silva

**Author notes:** equal contribution (Jaqueline Ramalho Buttura, Monize Nakamoto Provisor).

## Abstract

DNA mismatch repair deficiency (dMMR) leads to increased mutation load, which in turn may impact anti-tumor immune responses and treatment effectiveness. Currently, there are different mutational signatures described in primary cancers that are associated with dMMR. Whether the somatic and epigenetic changes in MMR genes precede one or more dMMR signatures, and if so by which mechanism remains unknown. To investigate the relationship between these changes and dMMR signatures, we performed a *de novo* extraction of mutational signatures in a large cohort of 787 gastric cancer patients. We detected three dMMR-related signatures, one of which clearly discriminates tumors with *MLH1* gene silencing caused by hyper-methylation within its promoter (AUC = 98%). We then demonstrate that samples with the highest exposures to signature share features related to better prognosis, encompassing clinical and molecular aspects, as well as altered immune infiltrate composition, predictive of a better response to immune checkpoint inhibitors. Overall, our analysis explored the impact of modifications in MMR-related genes on shaping specific mutational signatures and we provide evidence that patient classification based on mutational signature exposure can identify a group of patients with a good prognosis and who are potentially good candidates for immunotherapy.

## 1. Introduction

Cancer results from the sequential accumulation of DNA alterations, including single nucleotide mutations [1] that arise from different endogenous or exogenous processes [2]. Distinct DNA-damaging processes leave characteristic nucleotide base-change footprints known as mutational signatures [3]. Previous studies [4] have extracted distinct mutational signatures by examining a large set of human cancer genomes and some of these have been reported in the COSMIC database (denoted hereafter as CS). This pan-cancer analysis revealed significant heterogeneity of operational mutational processes, that encompass mutation-triggering events as diverse as the off-target activity AID/APOBEC family of cytidine deaminases, the exposure to ultraviolet light, tobacco-smoking and the defective DNA mismatch repair [5, 6].

Collectively, the understanding of the mechanistic basis of mutational signatures, as well as to their etiology, may provide clues for cancer diagnosis and hold prognostic value [7]. For example, six mutational signatures have been associated with BRCA1/BRCA2 dysfunction, which most likely are predictive of response to treatment with PARP inhibitors [8]. Thus, homologous recombination repair (HRR)-deficiency features based on these signatures allowed the prediction of BRCAness in breast cancer patients with 98.7% sensitivity [8]. Additionally, given that nucleotide excision repair (NER) deficient tumors are more sensitive to certain treatments, somatic variations in the *ERCC2* gene, which encodes a key protein of the NER pathway, have also been linked with characteristic mutational signatures [9, 10]. Other mutational processes are associated with patients harboring biallelic *MUTYH* germline mutations [11], a finding that may indicate deficient base excision repair (BER). Such patients are eligible for genetic counseling [12] and might benefit from immunotherapy [13].

In addition to HRR, NER and BER repair pathways, another mechanism underlying oncogenic genomic variations, with important effects on anti-tumor immune responses occur in tumors with impaired DNA mismatch repair (MMR), which harbor elevated frequencies of single-nucleotide variants (SNVs) and exceptionally high indel rates [14]. Recent studies demonstrated that various MMR-deficient (dMMR) tumor types (gastrointestinal, glioblastoma, endometrial and prostate) are more responsive to programmed cell death protein 1 (PD1) immune checkpoint inhibitors as compared to MMR-proficient tumors [15, 16, 17]. A set of four mutational signatures (CS-6, CS-15, CS-20, and CS-26) have been associated with dMMR. Nevertheless, it is still unclear if somatic and epigenetic changes in MMR genes lead to one or more dMMR signatures.

In this study we investigated the significance of molecular events in MMRgenes that shape characteristic mutation signatures found in MMR-deficient gastric adenocarcinomas. The presence of these signatures was evaluated for their prognostic value in a cohort of 787 gastric cancer patients with publicly available data, including 439 patients from the TCGA, and validated in a second cohort composed of 170 gastric cancer patients [18]. We further investigated whether local tumor immune response and prognosis varied according to MMR-deficiency exposure load. The consequences of these appear to be predictive of the responsiveness to immune checkpoint blockade and may be used to support treatment strategies in the future.

## 2. Materials and Methods

### 2.1. Clinical and genomic data from public cohort

The non-redundant public cohorts assessed here contained clinical and molecular information of gastric adenocarcinoma samples provided by: i) The Cancer Genome Atlas (TCGA, N=439), ii) cBioPortal, N=226; iii) and International Cancer Genome Consortium (ICGC, N=122), totaling 787 patients (Supplementary Material Table S1). TCGA data was assessed on October 4^*th*^, 2018 and corresponds to the MC3 variant calling project, which is a comprehensive effort to detect consensus mutations and forms the basis of Pan-Cancer Atlas initiative [19]. cBioPortal and ICGC cohorts’ comprise Asian samples which were last assessed on January 9^*th*^, 2019. Raw reads from matched non-tumor exomes from TCGA dataset, encompassing the MMR genes were downloaded and used to detect the germline SNVs following the Genome Analysis Toolkits (GATK)s best practice for germiline alterations calling. We also used additional filters considering mutations with VAF (variant allele frequency) ≥ 0.3 and minimum depth coverage of 10 reads. Furthermore, *dbNSFP_MetaLR_rankscore* was used to filter out (≤ 0.6) the synonymous mutations. The methylation levels in the form of beta-values ranging from 0 to 1 were addressed for the TCGA cohort [20]. We then used the CpG sites in the promoter of MMR genes to detect those that were hypermethylated or hypomethylated. The baseline clinical features are summarized in Supplementary Material Table S2.

### 2.2. Clinical and genomic data from validation cohort

Patients in the validation cohort were prospectively enrolled in an institutional study to unveil the epidemiology and genomics of gastric adenocarcinomas in Brazil [18]. This study was approved by the local ethics committee and all participants provided written informed consent. An overview of the clinical characteristics of patients in the validation cohort is provided in Table S3. Genomic DNA from frozen tissues (n=165) was extracted with AllPrep DNA/RNA Mini Kit (Qiagen), QIASymphony THC 400 (Qiagen) or phenol/chloroform/isoamyl alcohol precipitation. gDNA from FFPE tissue (n=4) was extracted with RecoverAll Total Nucleic Acid Isolation Kit (Thermo Fisher), and there was one sample from gastric wash. Exome libraries were prepared using Agilent SureSelect V6 kit and sequenced using Illumina platforms (HiSeq4000, 100bp, n=33; Novaseq, 150bp, n=137 - pairedend reads for both). The raw sequencing data (.fastq files) were deposited in SRA (http://www.ncbi.nlm.nih.gov/sra) under accession number PRJNA505810.

For our local independent validation cohort, the somatic SNVs were called by using an in-house pipeline following the Broad Institute GATK Best Practices guidelines [21] as described [6]. Briefly, the raw reads were aligned using Burrows Wheeler Aligner (BWA-mem) with default settings to assembly GRCh38. Next, alignment files in SAM format were converted to BAM files, sorted and filtered to exclude reads with mapq score <15. The retained reads were processed using SAMtools (v1.9) and Picard (v3.8) (*https://broadinstitute.github.io/picard/*) respectively, which excludes low-quality reads and PCR duplicates. Finally, the somatic SNVs calling was performed for the whole exome data from analysis-ready BAM files using Mutect2 (v3.8) for tumor samples and further with a panel of 16 unmatched non-tumor leukocyte samples. Extensive filtering was applied to remove low mapping quality, as well as strand, position bias and OxoG oxidative artifacts. Furthemore, any residual germline mutations from the database of germline mutations of of gnomAD (https://gnomad.broadinstitute.org/) and Online Archive of Brazilian Mutations (ABraOM, available at http://abraom.ib.usp.br/) were removed.

### 2.3. Mutational signatures estimation

All somatic SNVs of the six classes (C>A, C>G, C>T, T>A, T>C and T>G) were mapped onto trinucleotide sequences by including the 5’ and 3’ neighboring base-contexts. Next, the SNV spectrum with 96 trinucleotide mutations types for all samples were loaded into signeR [22] to estimate the optimal number of mutational signatures, which is based on the median Bayesian Information Criterion (BIC) value. We next used the cosine similarity to compare the extracted *de novo* mutational signatures to those described in the COSMIC signatures (v2), considering cosine similarity > 0.7 as a measure of closeness to COSMIC signatures. Patients with higher exposure for a given signature (Exposure value greater or equal to the third quartile) were named as *high* and those with lower exposure values (Exposure value less than third quartile) were named as *low*.

### 2.4. Molecular features

We used the MSIseq [23] software for microsatellite instability (MSI) status prediction (MSI-H and Non-MSI-H) from whole exome data. Briefly, this software is based on four machine-learning frameworks, which requires a catalog of somatic SNVs and microindels of samples, a file containing the exact locations of mononucleotides (length ≥ 5) and microsatellites consisting of di, tri, and tetranucleotide repeats, as annotated in the simpleRepeats track and avaliable at *http://hgdownload.cse.ucsc.edu/goldenpath/hg19/database/*. MSIseq is available at The Comprehensive R Archive Network (CRAN).

Consistent with a method previously proposed by Chalmers et al. [24], the Tumor Mutational Burden (TMB) was calculated as the total number of mutations divided by the length of the target region in megabases.

The tumor heterogeneity estimation was performed using math.score (MATH) function from package maftools version 3.8 [25]. A higher MATH score indicates increased tumor heterogeneity.

Neoantigen count: The list of neoantigens available for 77 TCGA-STAD samples was extracted from The Cancer Immunome Atlas (TCIA) [26], assessed on June 17th, 2017 at *https://tcia.at/neoantigens*.

### 2.5. Statistical analyses

The baseline patient characteristics are expressed as absolute and relative frequencies for qualitative variables and as the mean ± standard deviation (SD) for quantitative variables. Mutational signature exposure and TMB were considered as continuous variables. The association between qualitative variables was evaluated by chi-squared test or Fishers exact test, as appropriate.

Overall survival functions were estimated by the Kaplan-Meier estimator and the log-rank test was used to compare the survival functions among groups (eg, patients with higher mutational signature exposure (S^*high*^) versus other (S^*low*^)). The Cox semiparametric proportional hazards model was fitted to the dataset to describe the relationship between overall survival and the main clinical features. Hazard ratio (HR) and 95% confidence intervals (95%CI) were calculated for all variables. A backward stepwise selection algorithm was applied, with different significance levels to enter (p=0.10) and remain (p=0.05) in the model. Variables were removed from the model if they were non-significant or acted as confounders (change in coefficient >20%). The proportional hazards assumption was assessed based on the Schoenfeld residuals [27]. There was evidence that covariates had a constant effect over time in all cases.

Multivariate analyses were performed considering the main clinical features (such as age, pathological stage, Lauren tumor subtype and ethnicity), previously associated with overall survival, and with exposures of mutational signatures associated with dMMR, besides molecular features TMB and MSI status. Forest plots were created based on the final multiple Cox regression model. Metastatic patients were excluded from these analyses. In addition, we fitted simple and multiple logistic regression models in order to assess the effect of S2, S4 and S5 exposures in the *MLH1* methylation. Overall performance, calibration, and the discriminatory power of the final multiple logistic regression model were assessed using the Brier score, the HosmerLemeshow goodness-of-fit test, and the area under the receiver operating characteristic (ROC) curve (AUC), respectively [28]. Besides, we assessed the goodness-of fit through a Q-Q plot. The significance level was fixed at 5% for all tests (two-sided). Statistical analysis was performed using R software (v3.5).

### 2.6. Mutational signatures in cell lines

The CRISPR-Cas9 knockout clones for *MLH1* were generated in human HAP1 cells using the following guide RNA (gRNA) sequence: 5 - AAGA-CAATGGCACCGGGATC - 3. Clonal populations with a frameshift mutation within *MLH1* were subsequently cultured for three months to allow for the accumulation of mutations during cellular division [29]. To identify mutations, genomic DNA was submitted to whole genome sequencing (WGS). *De novo* somatic mutations including substitutions, indels and rearrangements in subclones were obtained by removing all mutations seen in parental clones. Next, SNVs were mapped onto trinucleotide sequences by including the 5’ and 3’ neighboring base-contexts and then the level of samples’ exposure to previously found mutational signatures was estimated [22].

### 2.7. Significantly mutated genes and pathway analysis

To assess the impact of dMMR pathway on the genes throughout the genome, we searched for genes more frequently mutated than would be expected by chance [30]. The proper gene symbol annotation in MAF (Mutation Annotation Format) files was addressed by maftools (v.8) [25] (prepareMutSig function) and then loaded into online MutSigCV server (v.1.3.4) (*https://cloud.genepattern.org/gp/pages/index.jsf*). The oncoplots were built by using the significantly mutated genes from MutSigCV analysis. The significantly mutated genes associated to S4^*high*^ and S4^*low*^ group were entered into Gene Set Enrichment Analysis (GSEA) according to the Investigate gene sets function available at MSigDB (Molecular Signatures Database (v7.0), *http://software.broadinstitute.org/gsea/msigdb/index.jsp*). KEGG, REACTOME, GO biological process, oncogenic signatures (module C6) and immunologic signatures (module C7) were also considered to compute overlaps, and the top 20 gene sets with FDR q-value <0.05 were used to summarize these analysis.

### 2.8. Inflammatory infiltrate and immune aspects

We estimated the cellular composition from the bulk expression datasets (TCGA) by using two complementary approaches. For both analyses, FPKM (Fragments Per Kilobase Million) from 380 tumor TCGA-STAD samples were used as normalized gene expression profile, retrieved on January 22^*th*^, 2018. First, the CIBERSORT software based on the deconvolution method for characterizing cell composition of complex tissues from their gene expression profiles, was used [31]. CIBESORT takes advantage of a validated leukocyte gene signature matrix, termed LM22. This gene signature contains 547 genes that distinguish 22 human hematopoietic cell phenotypes, including seven T cell types, nave and memory B cells, plasma cells, natural killer (NK) cells, and myeloid subsets. Simultaneously, a recent technique based on gene set enrichment analysis (GSEA) termed as xCell [32] was used to infer 34 immune cell types. Herein, we used this method to confirm the findings by CIBERSORT.

CIBERSORT analysis was performed online using a public server (*http://cibersort.stanford.edu/*) for characterizing absolute and relative immune cell composition with 1000 permutations and disabled quantile normalization as set parameters. From the 380 TCGA-STAD tumor samples, 215 215 samples (56%) yielded data on infiltrating immune cells (p-value<0.05), which were considered for further analysis (50 samples as S4^*high*^ and 165 as S4^*low*^). We also used the second approach known as xCell to reinforce the findings when comparing S4^*high*^ and S4^*low*^ samples. xCell analysis was performed using the R package with default parameters (available at *https://github.com/dviraran/xCell*). In order to verify the immune effector response present in S4^*high*^ and S4^*low*^ samples, differential expression of key immunoregulatory/inflammatory or cytotoxic markers was also performed. The comparison of the groups in this section was performed by *Mann-Whitney U* Test with statistical significance set at p-value<0.05.

We also used the pre-processed immune subtypes previously described by Thorsson et al. [33] for TCGA samples (available for 103 in S4^*high*^ samples and 285 in S4^*low*^ which can be assessed in Table S2).

## 3. Results

### 3.1. Mutational signatures

Using signeR [22] analysis to estimate *de novo* mutation signatures across three gastric cancer cohorts, we identified seven (denoted hereafter as S[1-7]) mutational signatures (Figure 1A) which are related to signatures described in the COSMIC database by cosine similarity scores (Figure 1B). Signature 1 (S1) is associated with endogenous mutational processes initiated by spontaneous deamination of 5-methylcytosine (CS-1); Signatures S2, S4 and S5 are associated with defective DNA mismatch repair and/or microsatellite instability (CS-6/CS-15, CS-20 and CS-21/CS-26 respectively); Signature S3 is related with failure of DNA-double strand break repair by homologous recombination (CS-3); Signature S6 related to CS-17 with unknown etiology; and Signature S7 is associated with error-prone polymerase activity (POLE (DNA Polymerase Epsilon, Catalytic Subunit), CS-10).

**Figure 1:**
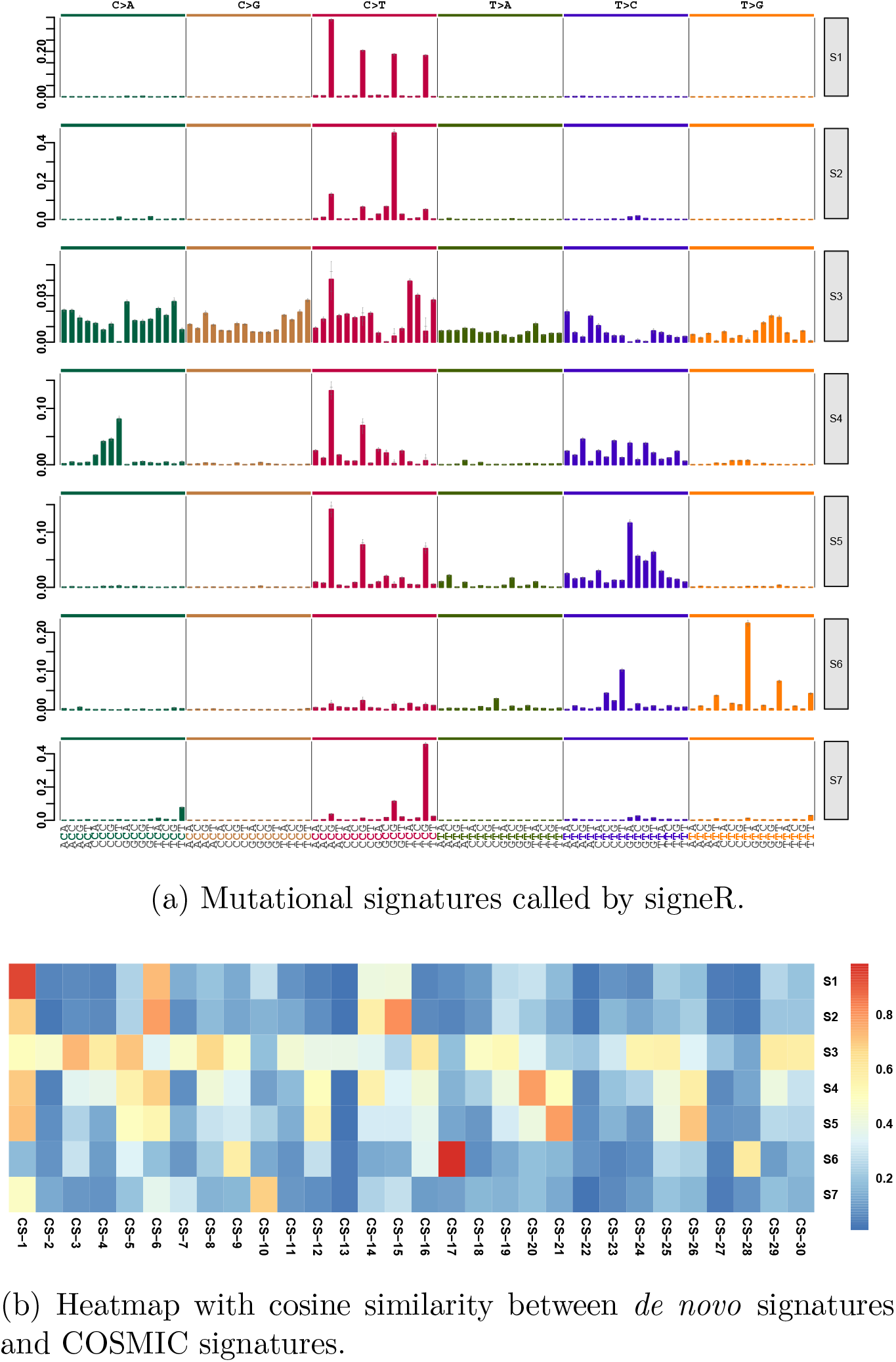
*De novo* mutational signatures in gastric cancer. (a) Mutational signatures in gastric cancer from 787 patients from TCGA, ICGC and cbioPortal cohorts. (b) Heatmap with cosine similarities between *de novo* mutational signatures and COSMIC signatures.

CS-3 (S3 in Figure 1 and Figure S1) was the predominant signature found here, supporting previous results which have characterized this signature in gastric cancer samples with a very high prevalence of small indels and base substitutions due to failure of DNA double-strand break repair by homologous recombination [34]. This finding suggests that 7-12% of gastric cancers may benefit from either platinum therapy or PARP inhibitors. However, notably, another group of patients not exposed to signature CS-3 was found to be highly exposed to signatures associated with dMMR (S2, S4 and S5 in Figure S1). Thus, our analysis identified a distinct group of gastric cancer patients harboring features that might have therapeutic relevance and which are further investigated here.

### 3.2. dMMR signatures and prognostic features

We reasoned that dMMR signature exposure could hold prognostic value in gastric cancer. Therefore, we first evaluated the influence of each dMMR signature exposure and the main possible clinical and molecular prognostic features such as age at diagnosis, ethnicity, tumor pathological stage, Lauren classification, anatomic site, TMB and microsatellite instability (MSI) status on overall survival (OS) fitting simple Cox regression model (Figure S2). Data from 584 gastric cancer patients with available vital status information (Alive/Dead) and without metastasis at diagnosis were included in simple and multiple Cox regression models. The median follow-up time for these patients was 28.9 months (with a 95% confidence interval: 95%CI 25.8-32.1) and the mean follow up time was 36.2 months (95%CI 32.9-39.5).

We then fitted a multiple Cox regression model to the dataset using prognostic features (variables with significant p-value are shown at Figure S2), and observed that S4 exposure burden was associated with improved OS compared with other dMMR signatures (hazard ratio [HR] 0.59 with 95%CI 0.37-0.96) (Figure 2A, Figure S3). Thus, we focused on signature S4, which has the potential to offer important clinically actionable information for treatment selection.

**Figure 2:**
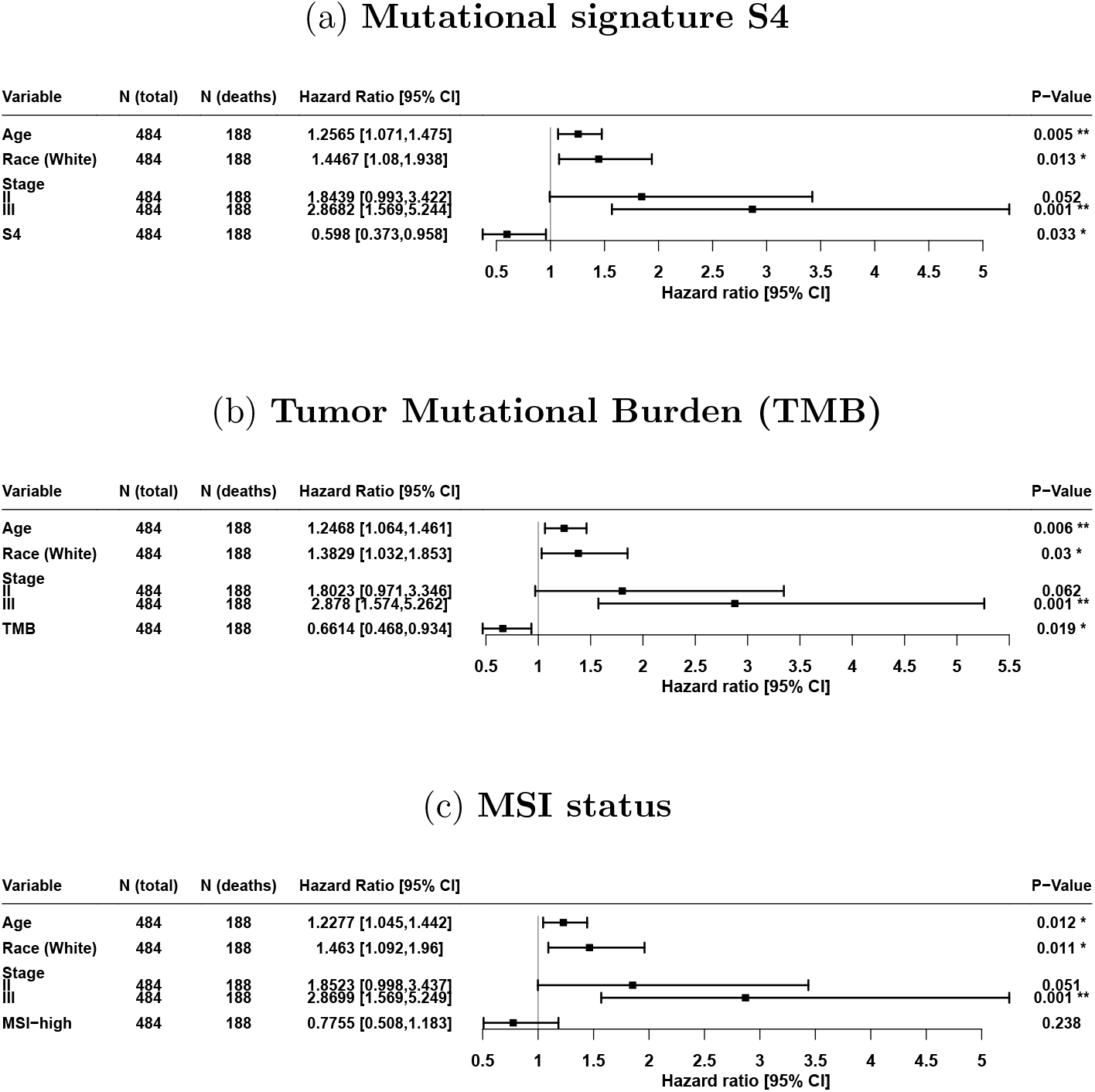
Forest plots showing the hazard ratio estimated according to multiple Cox regression models for overall survival.

Our analysis also revealed that a higher TMB was also associated with improved OS (HR=0.66; 95%CI 0.46-0.93) (Figure 2B), consistent with previous studies [35]. Distinctly from TMB, we found no association between predicted MSI-H status and improved OS (HR=0.78; 95%CI 0.51-1.18) (Figure 2C). The calibration curves for these models considering overall survival in 2 years is given in Figure S4, which indicates that all models are adequate.

We used the *maxstat* function (available in R language) to define groups according to the S4 exposure. The optimal cutpoint was within the range of highest quartile (Q3), and thus patients with S4 exposure ≥ Q3 were labeled as S4^*high*^ otherwise S4^*low*^ (<Q3). The survival curves from patients in the S4^*high*^ and S4^*low*^ groups were statistically different (p-value<0.03) with a median OS of 72 months (95%CI 48.0∞) in the S4^*high*^ group as compared to 37 months (95%CI 28.0-68.0) in the S4^*low*^ group (Figure S5). Next, we used an independent gastric cancer cohort to validate that S4^*high*^ has a survival benefit. Kaplan-Meier was performed to analyze patients in the validation cohort grouped based on samples’ exposure level of signature S4. By comparing patients in the S4^*high*^ group whose median OS was not reached at the time of 5 years (95%CI 38.2-∞) and the S4^*low*^ group with a median OS of 48 months (95%CI 21.3-∞), the data indicated a trend toward a survival benefit for S4^*high*^ group, supporting our previous findings (Figure S6).

### 3.3. dMMR signatures associated with MLH1 hypermethylation

Although the genes associated with dMMR are known, the underlying gene modifications that lead to each of the dMMR signatures still remain poorly characterized.

To improve our understanding of the determinant changes that influence the different dMMR signatures detected in this study, we first looked for somatic and germline SNVs and indels in MMR genes (*LIG1, POLE, EXO1, MLH1, MLH3, MSH2, MSH3, MSH5, MSH6, PCNA, PMS1, PMS2, PMS2L3, PMS2L4, POLD1, POLD2, POLD3, POLD4* and *SSBP1*). We observed that only 6% of patients (12/197) harbored somatic variations in the *MLH1* gene within either S2^*high*^ or S4^*high*^ groups and no mutated patients within S^*low*^ groups. Likewise, our results showed that 9% of the patients in the S2^*high*^ group harbored somatic variations in the *MLH3* gene. We also found that only 8% of patients in the S5^*high*^ group (8/100 considering TCGA cohort) harbor germline mutations in the *MSH5* gene. Altogether, we could observe only few cases harboring mutated MMR genes with some association with the S^*high*^ groups.

We next searched for epigenetic changes in the MMR genes. In line with previous studies [36, 37], we observed downregulation of *MLH1* gene expression driven by hypermethylation of its promoter (Figure S7). To further assess how the mutational exposure is associated with epigenetic changes in the *MLH1* gene, simple and multiple logistic regression models were fitted to the dataset (Table 1). This analysis revealed that S4 exposure burden was associated with an increased chance of *MLH1* promoter being methylated (odds ratio [OR]=22.561; 95%)CI 7.909-64.353). On the other hand, S5 exposure burden was associated with a decreased chance of *MLH1* promoter methylation (OR=0.107; 95%CI 0.048-0.238).Finally, no difference was observed in S2 exposure burden (OR=3.682; 95%CI 0.881-15.386). The performance of this model was adequate (HosmerLemeshow goodness-of-fit test *χ*^2^(8)=10.257; p-value=0.247) (Figure 3A), with a good performance observed (Brier score 0.0364), and an excellent power of discrimination (AUC=0.982; 95%CI 0.971-0.994) (Figure 3B). Using the Youden index, the best cutoff value (threshold) was 0.125, which had a sensitivity of 95.45% and specificity of 95.82% (Figure 3B).

**Table 1:** Simple and multiple logistic regression models for *MLH1* methylation and dMMR mutational signatures

| Variable | Simple logistic regression model |  |  |  |  |  |  |  |
| --- | --- | --- | --- | --- | --- | --- | --- | --- |
|  | Coefficient | Standard Error | CI(95%) for coefficient |  | p-value | OR | CI(95%) for OR |  |
|  |  |  | Lower | Upper |  |  | Lower | Upper |
| ExpS2 | 4.772 | 0.5226 | 3.748 | 5.796 | < 0.0001 | 118.155 | 42.424 | 329.078 |
| ExpS4 | 3.2424 | 0.3469 | 2.562 | 3.922 | < 0.0001 | 25.595 | 12.968 | 50.518 |
| ExpS5 | 0.2725 | 0.1674 | -0.056 | 0.601 | 0.104 | 1.313 | 0.946 | 1.823 |

| Variable | Multiple logistic regression model |  |  |  |  |  |  |  |
| --- | --- | --- | --- | --- | --- | --- | --- | --- |
|  | Coefficient | Standard Error | CI(95%) for coefficient |  | p-value | OR | CI(95%) for OR |  |
|  |  |  | Lower | Upper |  |  | Lower | Upper |
| Intercept | -2.640 | 0.300 | -3.227 | -2.052 | < 0.0001 |  |  |  |
| ExpS2 | 1.304 | 0.730 | -0.126 | 2.733 | 0.074 | 3.682 | 0.881 | 15.386 |
| ExpS4 | 3.1162 | 0.5348 | 2.068 | 4.164 | < 0.0001 | 22.561 | 7.909 | 64.353 |
| ExpS5 | -2.231 | 0.4067 | -3.028 | -1.434 | < 0.0001 | 0.107 | 0.048 | 0.238 |

**Figure 3:**
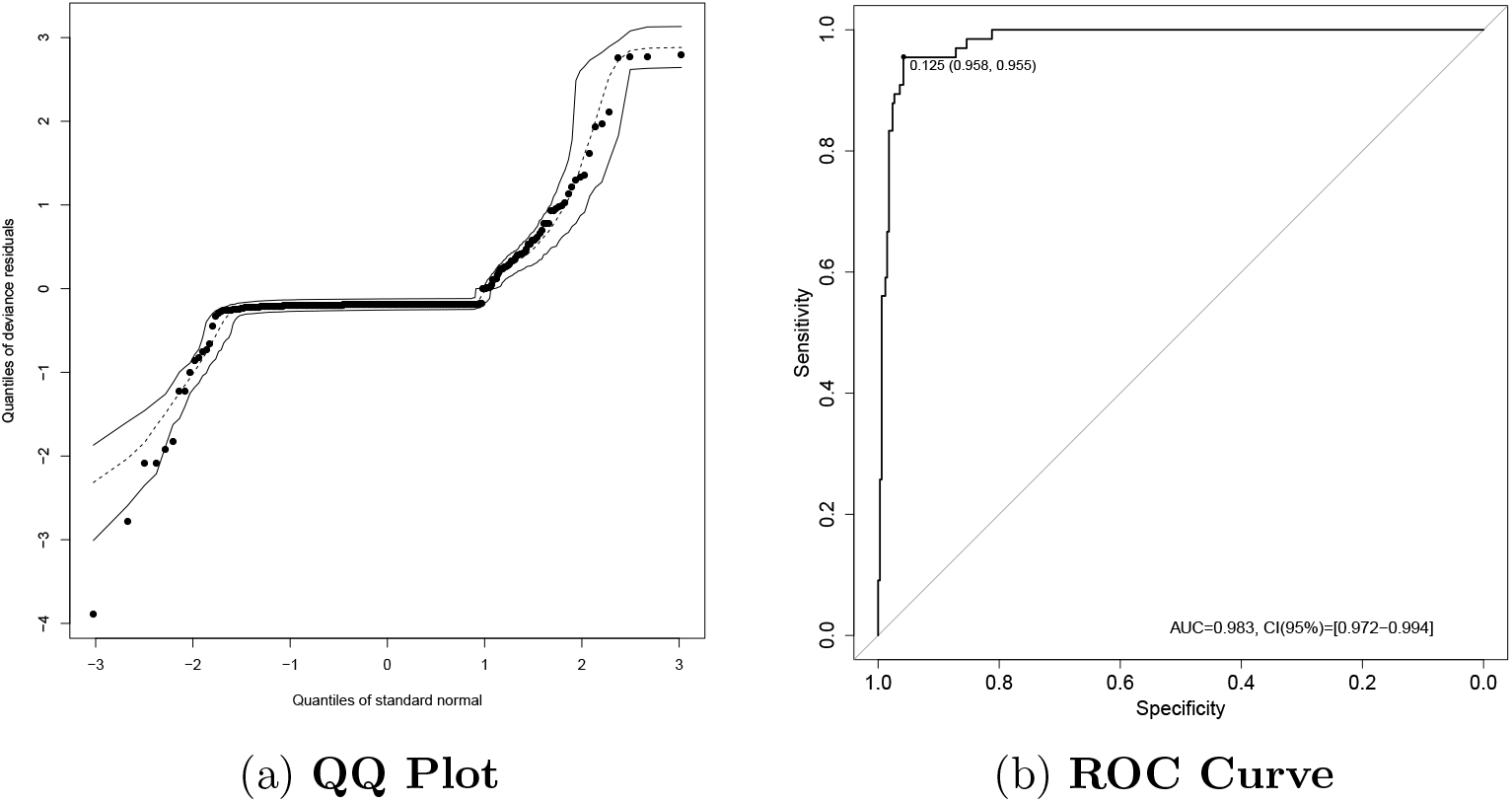
Performance and power discrimination of logistic model. (a) QQ Plot showing that the propensity score model had an adequate level of calibration according to the Hosmer-Lemeshow test. (b) Summary of ROC curve showing the power of discrimination for *MLH1* methylation at signature S4.

It should be noted that these observations suggest that neither germline SNVs, somatic SNVs or indels are the major modifications affecting gene expression levels, moreover, in fact, *MLH1* promoter is hypermethylated in almost 60% of individuals in the S4^*high*^ group (beta-value ≥ 0.3). Taken together, we conclude that the main mechanism of impaired MMR associated with the signature S4 (CS-20) in gastric cancer samples is driven by *MLH1* promoter hypermethylation. Moreover, by using genomic sequencing data from three HAP1 cells samples (2 *MLH1* ^*KO*^ and 1 *MLH1* ^*W T*^ cell), we have observed that *MLH1* ^*KO*^ cell lines have higher exposures of the S4 signature, while the parental cell line has higher exposure of the S5 signature (Figure S8), which identifies the absence of MLH1 as the cause of the S4 mutational signature.

### 3.4. Clinical and molecular features

Given our observation that the dMMR signature S4 was associated with better prognosis, and possibly related to an epigenetic causative mechanism, we next tried to further characterize the clinical and molecular features within S4^*high*^ and S4^*low*^ groups.

Previously defined clinical features that were associated with improved prognosis in gastric cancer were also enriched in the S4^*high*^ group (Table 2), such as distal anatomic site and intestinal histology [38]. On the other hand, known clinical variables associated with a worse prognosis in gastric cancer, such as cardia/proximal anatomic site, diffuse histology, positive lymph node metastasis (stage N+) and advanced pathological stages (stage III and IV) [38] were significantly higher in the S4^*low*^ group (Table 2). In addition, the predicted MSI-H status, MSI and POLE molecular subtypes were also enriched in the S4^*high*^ group, while genomically stable (GS) and chromosomal instability (CIN) molecular subtypes were enriched in the S4^*low*^ group (Table 2). Our data also reveal that most cases of MSI-H (n=119/160, 74%, Table 2) were grouped within the S4^*high*^ group, however, it has not escaped from our attention that a smaller fraction of MSI-H cases were unexpectedly grouped in the S4^*low*^ group. Similarly, we have also found Non-MSI-H patients in the S4^*high*^ group. Comparing the survival curves of these groups, we found MSI-H within S4^*low*^ group trends to have a worse prognosis with 9.07 months as median OS (95%CI 9.0-∞) than Non-MSI-H within S4^*high*^ group with 53 months as median OS (95%CI 20.0-∞). Similarly, diffuse histologic subtype grouped into S4^*high*^ (median OS not reached, 95%CI 24.0 *-*∞) trends to have a better prognosis than intestinal histologic subtype grouped into S4^*low*^ (median OS of 43.1 months, 95%CI 28.0-∞) (Figure S9). Thus, we conclude that the mutational signature classification was able to improve the stratification of patients within the prognostic groups, independent of their previous clinical or molecular classification.

**Table 2:** The clinical-pathological features of gastric cancer according to S4 dMMR mutational signature groups

| | $All(n = 787)$ | | $S4^{low}(n = 590)$ | | $S4^{high}(n = 197)$ | | $Pvalue$ |
| --- | --- | --- | --- | --- | --- | --- | --- |
| | $N$ | % | $N$ | % | $N$ | % | |
| **Age (mean ± SD)** | 64.17 ± 11.73 | | 63.44 ± 12 | | 66.25 ± 10.68 | | **0.0024** |
| **Gender** | 767 | 97 | 571 | 91 | 196 | 99 |  |
| *Female* | 274 | 36 | 192 | 34 | 82 | 42 | **0.0473** |
| *Male* | 493 | 64 | 379 | 66 | 114 | 58 |  |
| **Race** | 726 | 92 | 546 | 93 | 180 | 91 |  |
| *White* | 275 | 28 | 203 | 37 | 72 | 40 | 0.7427 |
| *Black* | 13 | 2 | 11 | 2 | 2 | 1 |  |
| *Asian* | 437 | 60 | 331 | 61 | 106 | 59 |  |
| *Other* | 1 | 0 | 1 | 0 | 0 | 0 |  |
| **Anatomic Site** | 627 | 80 | 495 | 84 | 132 | 67 |  |
| *Cardia/Proximal* | 168 | 27 | 148 | 30 | 20 | 15 | **0.0015** |
| *Fundus/Body* | 212 | 34 | 166 | 34 | 46 | 35 |  |
| *Antrum/Distal* | 242 | 39 | 178 | 36 | 64 | 48 |  |
| *Other* | 5 | 1 | 3 | 1 | 2 | 2 |  |
| **Histology Lauren** | 467 | 59 | 383 | 65 | 84 | 43 |  |
| *Diffuse* | 150 | 32 | 132 | 34 | 18 | 21 | **0.0041** |
| *Intestinal* | 301 | 64 | 235 | 61 | 66 | 79 |  |
| *Mixed* | 16 | 3 | 116 | 4 | 0 | 0 |  |
| **Stage T** | 712 | 90 | 526 | 89 | 186 | 94 |  |
| *T1 – T2* | 181 | 25 | 132 | 25 | 49 | 26 | 0.8116 |
| *T3 – T4* | 531 | 75 | 394 | 75 | 137 | 74 |  |
| **Stage N** | 712 | 90 | 526 | 89 | 186 | 94 |  |
| *N0* | 173 | 24 | 115 | 22 | 58 | 31 | **0.0144** |
| *N+* | 539 | 76 | 411 | 78 | 128 | 69 |  |
| **Stage M** | 707 | 90 | 524 | 89 | 183 | 93 |  |
| *M0* | 623 | 88 | 461 | 88 | 162 | 89 | 0.9422 |
| *M1* | 62 | 1 | 47 | 9 | 15 | 8 |  |
| *MX* | 22 | 3 | 16 | 3 | 6 | 3 |  |
| **Pathological Stage** | 715 | 91 | 546 | 93 | 169 | 86 |  |
| *I* | 85 | 12 | 58 | 11 | 27 | 16 | **0.0386** |
| *II* | 220 | 31 | 160 | 29 | 60 | 36 |  |
| *III* | 289 | 40 | 228 | 42 | 61 | 36 |  |
| *IV* | 121 | 15 | 100 | 18 | 21 | 12 |  |
| **Molecular Subtype** | 403 | 51 | 289 | 49 | 114 | 58 |  |
| *CIN* | 223 | 55 | 206 | 71 | 17 | 15 | <0.0001 |
| *GS* | 50 | 12 | 47 | 16 | 3 | 3 |  |
| *EBV* | 38 | 9 | 33 | 11 | 5 | 4 |  |
| *MSI* | 85 | 21 | 0 | 0 | 85 | 75 |  |
| *POLE* | 7 | 2 | 3 | 1 | 4 | 4 |  |
| **MSIseq Status** | 787 | 100 | 590 | 100 | 197 | 100 |  |
| *MSI – H* | 160 | 20 | 41 | 7 | 119 | 60 | <0.0001 |
| *Non – MSI – H* | 627 | 80 | 549 | 93 | 78 | 40 |  |
| **Immune Subtype** | 388 | 49 | 285 | 48 | 103 | 52 |  |
| *C1* | 128 | 33 | 107 | 35 | 27 | 26 | <0.0001 |
| *C2* | 209 | 54 | 135 | 47 | 74 | 72 |  |
| *C3* | 35 | 9 | 34 | 12 | 1 | 1 |  |
| *C4* | 9 | 2 | 8 | 3 | 1 | 1 |  |
| *C6* | 7 | 2 | 7 | 2 | 0 | 0 |  |

To further understand tumor heterogeneity in the S4^*high*^ and S4^*low*^ patients, we examined the spread of allele frequencies according to the quantitative measure of the degree of heterogeneity [39]. We then performed a correlation analysis based on this score, S4 exposure and TMB (Figure 4). We noted that the correlation of the tumor heterogeneity score (MATH) with either TMB or S4 exposure are opposite in the S4^*high*^ and S4^*low*^ groups. In the S4^*high*^ group, there was a negative correlation of S4 exposure or TMB with MATH and, in the S4^*low*^ group there was a positive correlation. We also observed that the MATH score is higher in the S4^*low*^ than the S4^*high*^ group (p-value=3.711×10^*-*12^). Lastly, the TMB and neoantigen load (by TCIA) have shown a positive correlation with signature S4 exposure in both groups (Figure 4).

**Figure 4:**
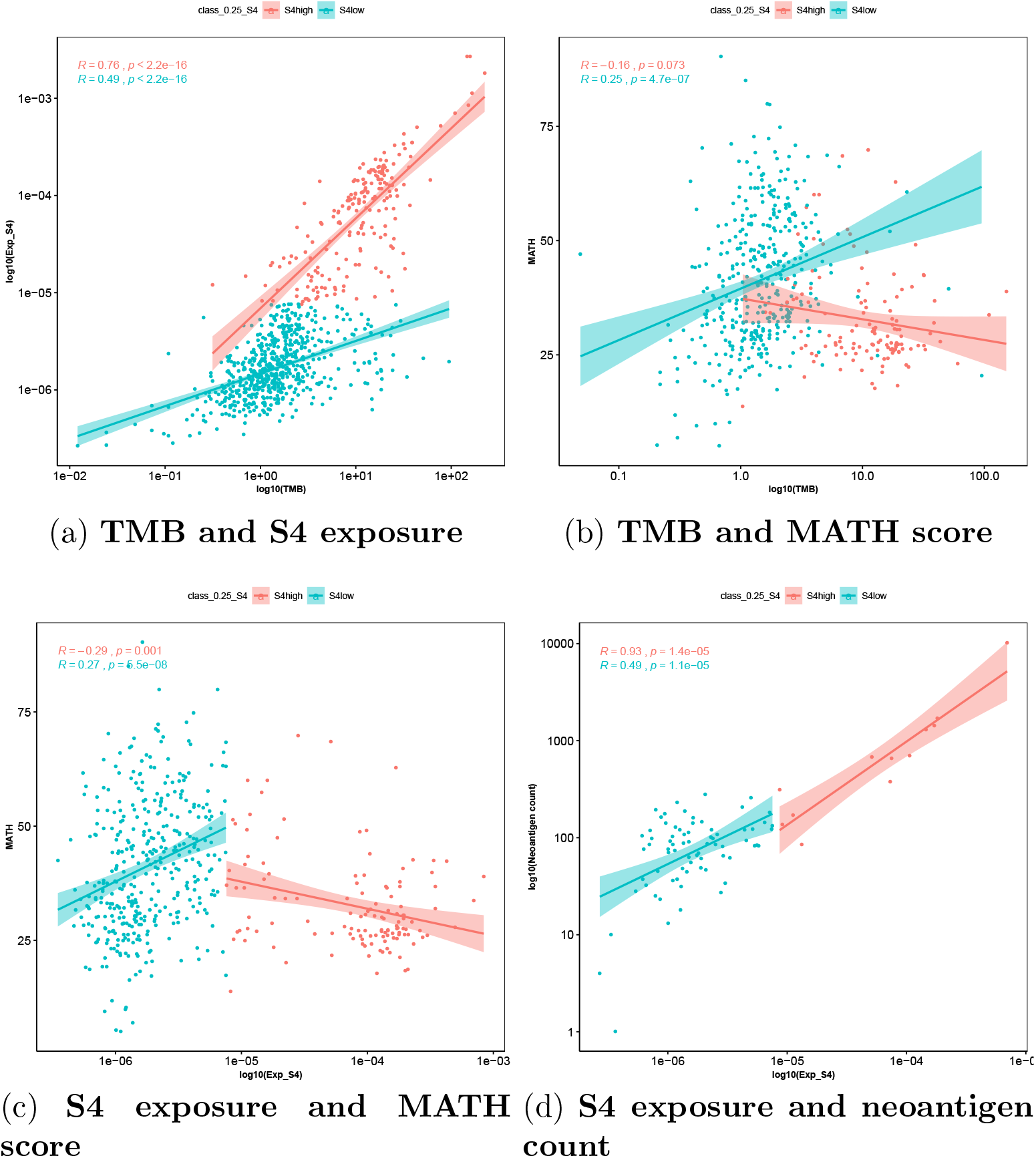
Scatter plots showing the Spearman correlation between molecular features. Blue dots represent patients in S4^*low*^ group and red those in S4^*high*^ group.

These findings suggest that tumors highly exposed to signature S4 are more homogeneous in the S4^*high*^ group and, together with high TMB and high neoantigen load, a reduced tumor heterogeneity appears to be determinant of a good prognosis. In this sense, we speculate that the methylation of *MLH1* promoter associated with the S4^*high*^ signature may be an early event in tumorigenesis.

In order to check if signature S4 is represented equally across the three cohorts, we compared their samples exposures. To avoid performing statistical tests with different numbers of samples, a subsampling procedure was applied, randomly selecting 24 samples from each cohort and then performing the KruskalWallis test. This was repeated 1000 times, always generating p-values above 0.05, leading us to conclude that the S4 exposure is similar for all cohorts.

### 3.5. Significantly mutated genes and related pathways in S4 dMMR groups

MMR-deficiency leads to an elevated frequency of mutations in the genome [14] and the consequences of MMR-deficiency may be derived from functional alterations in many distinct genes. In order to verify the existence of a common set of genes commonly mutated and their main related pathways between S4^*high*^ and S4^*low*^ groups (Supplementary Material Table S4) we investigated the presence of consistent SNVs differentiating these groups. At least one somatic mutation, including SNVs and indels, was detected for 83.25% of S4^*high*^ patients, and 78.64% within the S4^*low*^ group. We observed an increased number of deletions in the S4^*high*^ group, while within the S4^*low*^ group the mutations basically consisted of SNVs. These results are expected when considering that MSI/dMMR would lead to a higher number of deletions [14].

The gene set found as significantly mutated in the S4^*high*^ group is composed of 102 genes. The most commonly mutated genes in this group are *ARID1A* (42%), *KMT2D* (35%) and *TP53* (31%). In addition, there are another 56 genes presenting mutations in at least 10% of patients (Table S4A). The enrichment analysis of these mutated genes identified pathways related to immune cell differentiation, protein and RNA metabolism, gene expression regulation, cell differentiation and embryogenesis (Table S4B). It was previously suggested that somatic mutations in chromatinregulating genes such as *KMT2D* (also known as *MLL2*) and *ARID1A* are associated with improved survival [37].

In the gene set found as significantly mutated in theS4^*low*^ group, 12 out of 24 genes are known oncogenes, associated with tumor progression, or tumor suppressor genes. These 12 genes are *PIK3CA, KRAS, RHOA, CDH1, CTNNB1, ITGAV, SMAD4, TP53, CDKN2A, APC, PTEN* and *PIK3R1* (details in the Table S4C). The most frequently mutated genes in the S4^*low*^ group are *TP53* (47%), *ARID1A* (13%) and *CDH1* (9%) (Table S4C). The other 21 significantly mutated genes for this group were mutated in up to 8% of patients. In addition to the common pathways related to cancer, we also found pathways associated with regulation of cell death, phosphorus metabolism, regulation of transferase activity, morphogenesis pathways (gland development and anatomical structure of a tube) and VEGF and neurotrophin signaling pathways (Table S4D).

These findings were in accordance with some genes found previously in 215 non-hypermutated tumors from the TCGA cohort as *APC, CTNNB1, SMAD4* and *SMAD2*, with somatic mutations in *CDH1* and *RHOA* enriched in the genomically stable and/or diffuse histology [40], subtypes enriched in S4^*low*^ group.

### 3.6. Immune diversity in S4 dMMR groups

To investigate a possible role of the immune system being associated with the improved clinical outcomes seen in the S4^*high*^ as compared to S4^*low*^, we performed a series of analysis to determine the immune cell infiltrate composition in each group. These analyses used two different analytical methodologies (see Materials and Methods), and demonstrated a significantly higher proportion of infiltrating cytotoxic and pro-inflammatory immune cells in the group S4^*high*^,as exemplified by increased CD8+ central and effector memory T cells, CD4+ memory T cells, Th1 cells, gamma/delta T cells, NK cells, M1 macrophages and plasmacytoid dendritic cells (pDC), as compared to the S4^*low*^ group (Figure 5A and S10). In contrast, immature and immune regulatory dendritic cells were higher in the S4^*low*^ group (Figure 5A and S10).

**Figure 5:**
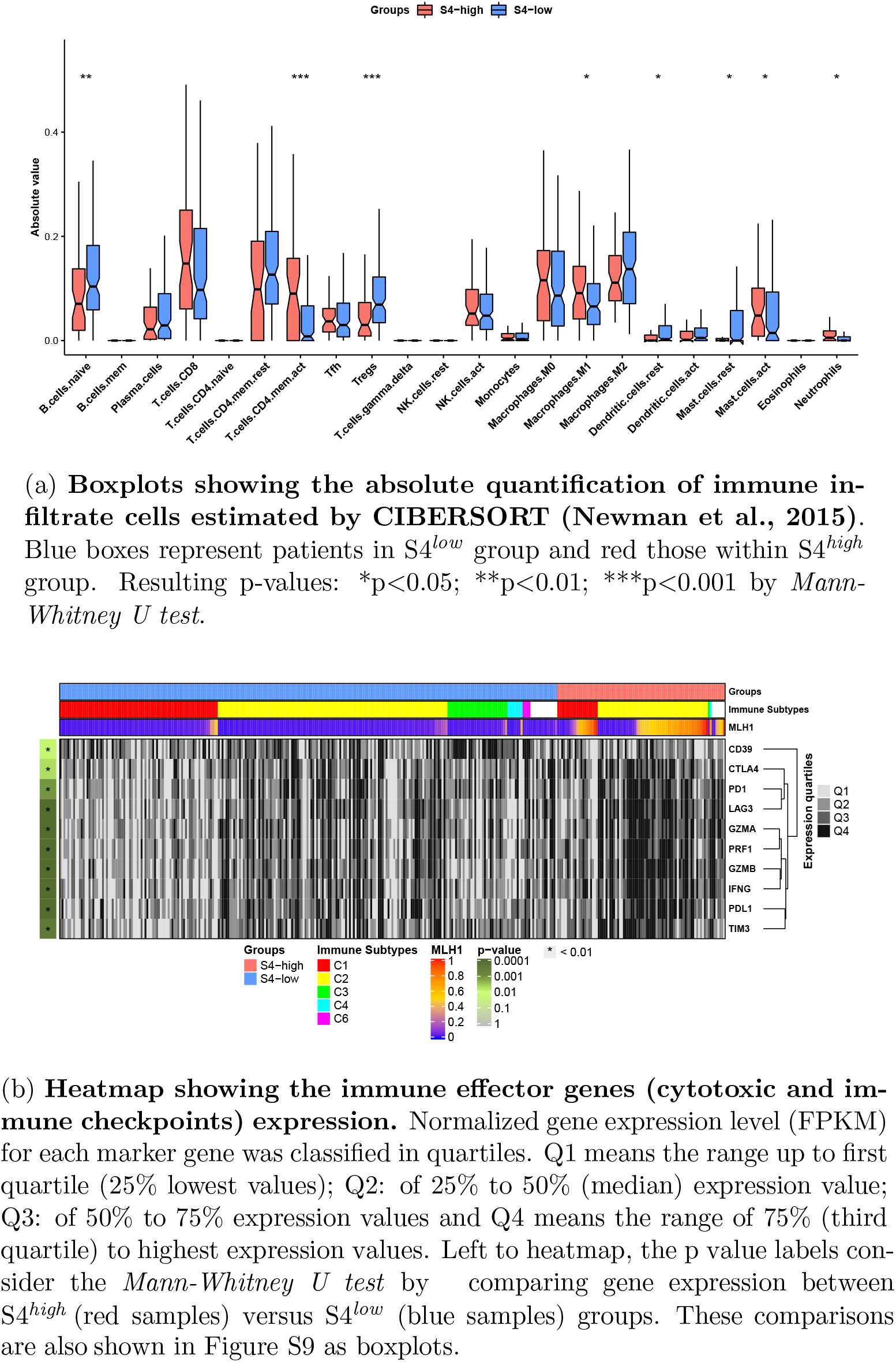
The main immunological features associated with S4^*high*^ and S4^*low*^ groups.

To further characterize the immune regulatory environment in the S4^*high*^ and S4^*low*^ groups respectively associated with good or poor clinical outcomes, we performed comparisons between gene expression levels of key genes coding for immunoregulatory and effector molecules proven to be important for tumor control in many cancers [41, 42]. First, the genes for CD8+ T cell related cytolytic molecules Granzyme A/B and Perforin-1 (*GZMA, GZMB* and *PRF1* genes, respectively) all displayed higher expression in the S4^*high*^ group (Figures 5B and S11). Moreover, the inflammatory T cell response related cytokine, IFN-gamma (*IFNG* gene) and other proinflammatory cytokines (*IL1B, IL6* and *IL8*), T cell activation marker genes (*IL2RA* and *ICOS*) and NK cell KIR family receptors were also higher in the S4^*high*^ than the S4^*low*^ group (Figures 5B and S11). Second, a series immunosuppression related genes (*TGFB1, IL10*, and *FOXP3*) showed no differences between the groups, while *ENTPD1* (CD39 gene), a protein associated with Treg immunosuppression activity [43], was higher in S4^*low*^ vs. S4^*high*^ (Figures 5B and S11).

Importantly, the expression of the immune checkpoint inhibitor genes (*PDCD1* for PD1 receptor, *CD274* for PD-L1 ligand, *PDCD1LG2* for PD-L2, *HAVCR2* for TIM3, *LAG3* and *CTLA4*) were also higher in the S4^*high*^ group (Figures 5B and S11) possibly indicating a relationship with a more immunologically activated tumor microenvironment [44]. Expression of *HLA*, antigen processing and presentation-related genes (such as *CD86, B2M*, various *HLA* class II genes, *HLA-E, HLA-C, TAP1* and *TAP2*) were also higher in the S4^*high*^ group (Figure S11). Together these findings indicate a highly activated immune microenvironment in the S4^*high*^ group as compared to the S4^*low*^ group.

The immune subtypes previously characterized by Thorsson et al. [33] reinforce the finding that the S4^*high*^ group primarily presents a more immunologically active tumor microenvironment which is composed predominantly of C2 (interferon-gamma) immune subtype, with a significantly higher proportion of this subtype as compared to S4^*low*^ (Table 2 and Figure 5B). Importantly, the C2 immune subtype has been associated with highly mutated tumors [33]. On the other hand, the S4^*low*^ group displayed a higher proportion of C3 (inflammatory) and C1 (wound healing) immune subtypes [33] (Table 2 and Figure 5B). Lastly, the C2 immune subtype in the S4^*low*^ group seems to be less activated than in the S4^*high*^ group with reduced relative gene expression of immune effector molecules (Figure 5B).

### 3.7. Discussion

This study provides a comprehensive and integrated analysis of the impact of MMR-related gene alterations in shaping specific mutational signatures associated with gastric cancer. We present evidence that the determination of MMR-deficiency can be used not only for MSI-phenotype classification, but also as a potential indicator of prognosis and to select potential candidates for treatment with checkpoint inhibitors.

We performed a *de novo* extraction of mutational signatures based on somatic SNVs across four WES cohorts, spanning 787 gastric cancer samples derived mainly from populations with European and Asian descendance. We found 7 different mutational signatures, with three elated to MMR-deficiency.

Next, we examined the prognostic value of these dMMR signatures in multivariate survival analysis by employing a Cox proportional hazards model. This analysis revealed signature S4, related to the previously described CS-20, as the only dMMR signature with significant prognostic value. This prognostic value was validated using our local cohort of gastric cancer patients, distinct in terms of molecular ancestry as well as some clinical and molecular features such as Lauren’s histology and tumor heterogeneity. This cohort was predominantly composed of diffuse/mixed histology samples, while public cohorts were enriched for the intestinal subtype. Furthermore, the S4^*low*^ group in this independent cohort was less heterogeneous than the S4^*low*^ group from the public cohorts and even than the S4^*high*^ groups from both cohorts. Nevertheless, we observed a better prognosis for the patients of the S4^*high*^ group, also for this cohort.

Interestingly, after performing a comprehensive analysis of patients exposed to signature S4 by an in depth evaluation of molecular and immune features, we observed that the main mechanism associated with impaired MMR seems to be the hypermethylation of the *MLH1* gene promoter (*hMLH1*). Moreover, we show that disruption of *MLH1 in vitro* using CRISPR/Cas9 assay reproduces the CS-20 signature [45] that resembles the S4 signature. Here, we have shown an endogenous epigenetic mechanism for this signature in gastric cancer patients. Remarkably, we also reproduce the S4 signature in an isogenic cell model in which the MLH1^*KO*^ cells had a high exposure of the S4 signature. Thus importantly, we conclude that independently of the primary mechanism that leads to the loss of *MLH1* gene expression - due to promoter hypermethylation or loss of function mutagenesis - it results in the same mutational signature.

It has been well documented that CpG island methylator (CIMP) phenotype is an early event in tumorigenesis, preceding the *hMLH1* in solid tumors, which in turn drives the microsatellite instability high (MSI-H) phenotype [36, 40, 37]. In contrast, MSI-low (MSI-L) and microsatellite stable (MSS) gastric carcinoma subtypes have unmethylated *MLH1* promoters and regular MLH1 activity [36]. Here, we classified samples as MSI-H and non-MSI-H (MSI-L and MSS) and observed that most cases of MSI-H fall within the S4^*high*^ group, however, a smaller fraction of MSI-H cases did not show high exposure of this mutational signature and were grouped in the S4^*low*^ group. This is in line with previous studies showing about one-quarter of the MSI-H cases, despite being MSI-H, present distinct molecular features and poor prognosis [46]. Similarly, we have also found Non-MSI-H patients in the S4^*high*^ group, showing that mutational signature exposure is capable of clustering samples independently of their MSI-status. Furthermore, we also identified a few cases (4%) in the S4^*high*^ group that instead of presenting *hMLH1*, carried somatic mutations in the *MLH1* gene that apparently lead to loss-of-function of the encoded protein. Thus, for about 70% of S4^*high*^ cases we found a clear genetic or epigenetic cause.

We also demonstrated a strong correlation between S4 exposure and TMB, which showed significant prognostic value upon multivariate survival analysis. Hypermutated tumors have been associated with better prognosis and a good response to immunotherapy apparently due to neoantigen enrichment and intrinsic antitumor immune responses [47, 48]. However, a threshold for classifying TMB-high samples usually varies with tumor type [49] and in some cases may not predict a better response [48] due to intratumoral heterogeneity [50]. In this sense, it is important to highlight that most mutations in the S4^*high*^ signature are clonal, which is an important feature to predict response to immune checkpoint inhibitors therapy.

High intratumoral heterogeneity has been associated with an incomplete response to therapy, higher relapse rates, and poor clinical outcomes [51, 52]. The increased genomic instability observed in MSI/dMMR and CIN (chromosomal instability) tumors is the major driver of high intratumoral heterogeneity [53, 51]. However, the most unstable tumors (with the highest burden of somatic SNVs or copy number alterations) are not the most intrinsically heterogeneous[53]. Furthermore, the greatest intratumor heterogeneity was found in tumors exhibiting relatively high numbers of both somatic mutations and copy number alterations, which can be associated with exogenous mutagens, including viral infection and tobacco smoking. These tumors have high number of sub-clonal mutations related to late events and exhibit frequent chromosomal instability associated with CIN subtype, *TP53* mutations, and APOBEC-related mutational signatures (previous related to EBV gastric cancer subtype [6, 53]. Likewise, here we noted that patients with higher S4 exposure harbor more homogeneous tumors as compared to S4^*low*^ group. Taken together, the S4^*high*^ group encompassed patients with intermediate to high TMB, in addition to the MSI and CIN molecular phenotypes associated with lower tumor heterogeneity, which might allow for a more effective antitumor immune response in this subset of gastric cancer patients. In this sense, a recent meta-analysis discussed the importance of MSI-status for the treatment response in gastric cancer patients, suggesting that MSI-H patients may not benefit from perioperative or adjuvant therapy and could go straight to surgery [54].

Finally, several studies have shown that the tumor microenvironment context, at diagnosis, is capable of predicting treatment response and clinical outcome [55, 56]. The balance of inflammatory/cytotoxic immune cells, with elements of an effective antitumor response, including regulatory cells and suppressor signals, may indicate which patients have an intrinsically effective antitumor response, and thus, a better prognosis. EBV and MSI subtypes in gastric cancer have already been associated with higher immune infiltrate and responsiveness to immunotherapy, as well as better prognosis [55]. Here we found many elements indicating that the tumor microenvironment in the S4^*high*^ group is more active as compared to S4^*low*^ patients. In general, the absolute quantification by CIBERSORT [31] or GSEA scores by xCell [32] of immune cell subtypes and the differential gene expression pointed to higher activity of proinflammatory and cytotoxic cells, as well as antigen processing and presentation in S4^*high*^. In contrast, although there were some immunogenic tumors in S4^*low*^ group, the predominant environment was enriched in Treg lymphocytes and M2 macrophages, both related to worse prognosis [56, 55].

In conclusion, while past studies have aimed to identify patients using molecular and clinical features such as MSI status, TMB load and *MLH1* gene expression levels, our study provides evidence that classification based on mutational signature exposure may identify groups of patients with common clinical, immunological and mutational features that are directly related to a better prognosis, and who might benefit from immunotherapy-based treatments.

## Data Availability

The raw sequencing data (.fastq files) were deposited in SRA (http://www.ncbi.nlm.nih.gov/sra) under accession numbers PRJNA505810 and

## 3.8. Acknowledgements

This project received financial support from FAPESP (14-26897-0 and 16/11791-7); ED-N and KJG are research fellows from Conselho Nacional de Desenvolvimento Científico e Tecnolgico (CNPq, Brazil). ED-N acknowledges the support given by Associação Beneficente Alzira Denise Hertzog Silva (ABADHS).

## Notes

### Competing Interest Statement

The authors have declared no competing interest.

